## Supplementary materials for "Generalizing population RT-qPCR cycle threshold values-informed estimation of epidemiological dynamics: Impact of surveillance practices and pathogen variability"

### Supplementary methods

#### 1. Simulation and viral dynamics

##### *Simulation of consecutive epidemic waves*

We used a susceptible-exposed-infectious-recovered (SEIR) model to simulate two consecutive epidemic waves in a closed population of 7.5 million (similar to Hong Kong's population) with initial infections at 0.001%. The model was implemented using the R package odin(1), which employs stochastic differential equation solvers to capture variability in disease transmission. Briefly, we simulated populations with compartments for susceptible (S), exposed-but-not-yet-infectious (E), infectious (I) and recovered (R). The compartmental transition equations are listed below:

$$\begin{aligned} \frac{dS}{dt} &= \frac{-\beta_t S(t) I(t)}{N} \\ \frac{dE}{dt} &= \frac{\beta_t S(t) I(t)}{N} - \sigma E(t) \\ \frac{dI}{dt} &= \sigma E(t) - \gamma I(t) \\ \frac{dR}{dt} &= \gamma I(t) \end{aligned} \tag{1}$$

where $\beta_t = \frac{R_0}{\gamma}$ for $t \geq t_0$. $1/\sigma$ indicated the average time for individuals to transit from E to I (i.e. incubation period), while $1/\gamma$ referred to the observed mean infectious period and was fixed at 5 days(2). Detailed descriptions of parameters were listed in Table S1.

We used synthetic $\beta$ (which determines the underlying transmission rate) that changed over time $t$ to synthesize the process of two consecutive epidemic waves:

$$\beta_t = \begin{cases} R_0\gamma & t < 60 \\ R_0^2\gamma & 60 \leq t < 110 \\ R_0^3\gamma & 110 \leq t < 150 \\ R_0^4\gamma & t \geq 150 \end{cases} \tag{2}$$

where $R_0 = 2.2$, $R_0^3 = 1.9$ and $R_0^2 = R_0^4 = 0.3$ if otherwise specified. Epidemic switch points were set at day 60 and 110 and $R_0$ changes between switch points were fitted via a cubic smoothing spline and interpolated into smooth transitions. $R_t$ calculated under this SEIR model (denoted as the simulation truth) would then be:

$$R_t = \frac{S(t)}{N}\beta_t\gamma \tag{3}$$

***Individual viral load trajectories***

We simulated viral load trajectories over the course of infection for symptomatic cases using a previously published method(3, 4). Briefly, the cycle threshold (Ct) values of infected individuals change in a unimodal pattern, reaching the lowest point (peak viral load) at illness onset and returning to normal within 2-3 weeks after onset. The peak viral load ($VL_p$) follows a normal distribution, while the duration of viral shedding since onset ($T_s$) is lognormally distributed (Table S1).

In scenarios considering infection severity (i.e., scenarios 7-14), the duration of viral shedding since onset for severe, critical and fatal cases were assumed to be 2.3, 6.5 and 13.1 days longer than mild cases(5).

Each infected symptomatic individual has their simulated Ct values per day after infection, and their sampled Ct values correspond to the Ct value falling on day $k$ post infection based on their individual Ct trajectories, where $k$ is the interval between infection and detection.

***Aligning viral shedding durations to infection***

In our main analyses, viral load is assumed to peak at illness onset. To explore the impact of different timings of viral peaks, we updated the distribution of viral shedding duration to align with infection rather than symptom onset, which allows more flexible comparisons between pathogens.

We computed 1000 random values each for incubation periods and viral shedding durations since onset, based on the basic distributions used in the main scenarios (see Table S1). These two series of numbers were then summed, and the resulting 1000 joint values were used to fit the distribution of viral shedding duration since infection. We fitted these 1000 joint values to both normal and lognormal distributions and compared their Akaike Information Criteria (AIC). The lognormal distribution was chosen due to its lower AIC. The viral shedding duration since infection, as updated using this method, followed a lognormal distribution with a mean of 22.6 and standard deviation (SD) of 1.3.

***Shedding parameters for real-world pathogens***

We searched literature for parameters of incubation periods, viral shedding durations, quantities of viral peak and the timing of viral peak for different variants of SARS-CoV-2, SARS-CoV-1 and influenza A. We converted the incubation period distribution from available literature to lognormal distribution to allow more unified parameters for comparing across pathogens, which also enabled more efficient $R_t$ estimation using EpiNow2. As viral shedding durations were sometimes reported from different start points including onset or inoculation/infection, we used the approach mentioned above to derive the distribution of viral shedding durations since infection for consistency. For literature that only reported the confidence intervals (CIs) but not the SD for the incubation period, we approximated their SD by dividing the length of the CIs by 3.92 assuming normal distributions. The original and converted distribution of viral shedding durations for included pathogens is shown in Table S3.

Due to differences in viral load measures across studies and laboratory methodologies, we utilized the relative differences in viral peaks between pathogens to parameterize their viral peaks. For instance, the viral peak of the Alpha variant was found to be higher than that of the ancestral strain ($10^{7.7}$ vs $10^{7}$)(6). In this case, we first converted the Ct values into RNA copies using a formula from the literature(7), then applied the relative difference between the two pathogens to determine the peak viral load in RNA copies for the Alpha variant. Lastly, we converted the RNA copy quantities back into Ct values to approximate the distribution of the viral peak for Alpha.

The six selected pathogens exhibit different viral shedding characteristics (Fig 4A; Table S3). For instance, both Alpha and Delta variants of SARS-CoV-2 displayed higher viral peak compared to the ancestral strain, with both peaking around or before illness onset(6, 8). However, the shedding duration was shorter for the Alpha variant and longer for Delta(6, 9). On the contrary, despite having similar peak viral loads, the ancestral and Omicron strain of SARS-CoV-2, SARS-CoV-1 and influenza A showed considerable differences in their other shedding characteristics. The Omicron strain and SARS-CoV-1 exhibited later and much later viral peak respectively(10-14), while influenza A had a much shorter shedding period(15) (Fig 4A). It is important to note that there is controversial evidence regarding the timing of the viral peak relative to symptom onset for the Delta variant. In our analyses, we used the evidence suggesting that the viral peak occurs before onset(6) to provide an example of this specific relationship between viral peak and symptom onset.

***Exploring Ct testing window for Omicron scenario***

In our analyses, detected cases were typically tested for Ct values at the time of detection. To assess the impact of adjusting the Ct testing window for the Omicron scenario, we introduced a delay in Ct testing by 2, 3, or 4 days after case detection and assigned a detection-to-testing delay to detected individuals. Same to other scenarios, Ct values at sampling were used to reflect the Ct distributions for each day to estimate Ct-based $R_t$.

This adjustment allowed us to capture the monotonic shedding phase of Omicron infection, leading to improved model accuracy (Fig S9). This highlights the importance of monotonic shedding trajectories for accurate Ct-based $R_t$ estimation.

#### 2. Estimation of $R_t$ and evaluation of accuracy

***Estimation of incidence-based $R_t$***

Incidence-based $R_t$ from synthetic case count data was estimated using the R package EpiNow2(16), which has accounted for delays and other sources of uncertainty. The incubation period and reporting delay that were used for deconvolution were assumed to follow the delay distributions that were simulated from symptom-based surveillance. The mean and variance of the generation interval under the SEIR model was specified as $T_c = 1/\sigma + 1/\gamma$ and $\text{Var} = 2(\frac{T_c}{2})^2$ respectively, with $\sigma$ and $\gamma$ being the average time for individuals to transit from E to I and from I to loss of infectiousness respectively (Table S1). More details were provided in https://github.com/epiforecasts/EpiNow2.

***Optimal training periods for all scenarios***

To establish a generic training period applicable to all scenarios, we compared the adjusted R-squared values of regression models trained over different time periods with varying start dates and durations. We evaluated 40 alternative start dates, ranging from the first date when the case count exceeded 15 to 39 days onward. For the durations of these alternative training periods, we considered 30, 40, 50 and 60 days respectively.

The optimal training period was identified as the one that provided the highest adjusted R-squared among the 160 possible combinations of training periods within each scenario. Such optimal training periods typically aligned with the epidemic's peak and transition point (i.e., when incidence-based $R_t$ shifted from above 1 to below 1), covering the days with the most data (Fig S1). Therefore, we set the training period as three weeks before and after the date of highest cases count during the first epidemic wave (day 0-109 after the initial outbreak) to be applied to all scenarios.

##### *Additional evaluation metrics*

We included additional evaluation metrics to assess model performance across simulated surveillance scenarios. Spearman's correlation coefficient ($\rho$) was calculated to evaluate the consistency between Ct-based $R_t$ and the simulation truth, while mean absolute percentage error (MAPE) measured the average percentage deviation of estimates from true values(17):

$$\text{MAPE} = \frac{1}{D}\sum_{d=1}^{D} \left| \frac{\ln\left(R_{t,d}\right) - \ln\left(E\left(R_{t,d}\right)\right)}{\ln\left(R_{t,d}\right)} \right| \tag{4}$$

Here, $D$ is the number of days in the testing period, $E\left(R_{t,d}\right)$ is the estimated Ct-based $R_t$ and $R_{t,d}$ is the simulation truth on day $d$. Notably, MAPE is particularly useful for understanding relative error and comparing performance across scenarios with different scales, such as scenario 6 and others.

These metrics complement the area under the receiver operator characteristic curve (AUC), providing a comprehensive view of the model's estimation accuracy. Notably, scenarios 6, 10, and 14 consistently showed less favourable performance across all metrics (Fig S3), highlighting the robustness of the evaluation framework.

##### *Evaluate uncertainty in estimation using bootstrap sampling*

We used a non-parametric bootstrap method to assess the uncertainty in estimation accuracy of Ct-based $R_t$ as measured by AUC and other evaluation metrics. We repeated each scenario 100 times and resampled Ct values from cases that had their sampling on each day based on the daily sample count. This allowed us to reconstruct the population Ct values for each day. We then re-estimated the Ct-based $R_t$ using the updated population Ct values and calculated the evaluation metrics, including the AUC, by comparing the new Ct-based $R_t$ to the simulation truth. We obtained 100 values for each evaluation metrics through bootstrapping and summarized the estimation accuracy by taking the median, 2.5% and 97.5% quantiles for each scenario.

For scenarios with different case severity profiles (i.e., scenarios 7-14), we conducted resampling separately for mild cases and those with severe or above conditions. By counting the daily number of mild and severe cases and resampling accordingly, we ensured the proportion of severe cases detected per day remained consistent across iterations.

***Exploring the threshold for sufficient Ct value samples***

We explored estimation accuracies in relation to the number of Ct values available from detected cases per day to determine whether the sample size affected estimation accuracies. We randomly selected *N* Ct values from detected cases each day, with *N* varying from 10 to 200 (e.g., 10, 20 and 30). If fewer than N cases were detected on a given day, all available Ct values were used. Performance evaluation was skipped if the daily number of detected cases was consistently below *N* throughout the entire period.

This sampling strategy was repeated 100 times for each threshold *N.* In addition to comparing absolute estimation accuracies, we calculated the relative change in AUC between each two sample count thresholds to understand the incremental value of increasing the daily sample size. This was done by computing:

$$\Delta AUC_{i+1} = \frac{AUC_{i+1} - AUC_i}{AUC_i} \quad (5)$$

Where *i* is the *i*-th sample count threshold *N* and $AUC_i$ is the AUC at that threshold. The median, 2.5% and 97.5% quantiles of the AUC and $\Delta AUC$ for different sample count thresholds were used as summary statistics.

By exploring the threshold for sufficient Ct value samples, we found that estimation accuracies improved with more samples, but the improvement became marginal beyond 100 samples, with ΔAUC remaining below 2% and overlapping with the AUC for sampling all cases (Fig S6).

#### Supplementary tables

**Table S1. Parameters for simulating consecutive epidemic waves using the SEIR model.**

| | Name | Description | Values |
|---|---|---|---|
| **Epidemiological parameters** | $R_0$ | Basic reproductive number. | 2.2, 0.3, 1.9 and 0.3 for two typical waves; 2.2, 0.5, 1.1 and 0.9 for a typical first wave and a flat second wave |
| | N | Initial population size. | 7,500,000 (population size in Hong Kong) |
| | $I_0$ | Proportion of individuals infected at seeding time. | 0.001% |
| | $1/\gamma$ | Average time for individuals to transit from infectious ($I$) to loss of infectiousness (mean infectious period). | 5 days(2) |
| | $1/\sigma$ | The average time for individuals to transit from exposed-but-not-yet-infectious ($E$) to infectious ($I$). | 5 days |
| | $T_{incu}$ | Incubation period. | lognormal (mean=5.2, SD=3.9(18)) |
| | $p_{Symp\|Inf}$ | Probability of individuals who developed symptoms after infections. | binomial (0.6(3, 19)) |
| | $p_{Detect\_s\|Symp}$ | Probability of detection under symptom-based reporting. | 25%, SD=0.5% in a typical scenario (i.e. scenario 1) but could vary in different scenarios |
| | $T_{Detect\_s}$ | Onset-to-detection delay (in days). | gamma (shape=1.83, rate=0.43) |
| **Viral kinetics parameters** | $VL_p$ | Peak viral load. | normal (mean=22.3, SD=4.2(20)) |
| | $T_s$ | Time from onset to cessation of viral shedding. | normal (mean=17, SD=0.79(10)) |

**Table S2. Detailed descriptions of simulated scenarios to explore the impact of surveillance coverage.**

| | Characteristics | | | |
|---|---|---|---|---|
| **Scenario** | **Epidemic** | **Case detection** | **Detection delay** | **Ct testing** |
| 1 | Two typical waves# | Symptomatic cases have a 25% detection probability during both waves. | Similar detection delays for all cases. | All detected cases will be tested with Ct values. |
| 2 | | | | 80% of detected cases will be tested with Ct values. |
| 3 | | | | 50% of detected cases will be tested with Ct values. |
| 4 | | | | 30% of detected cases will be tested with Ct values. |
| 5 | | Symptomatic cases have a 25% detection probability during the first wave;<br>only around 100 cases per day will be detected over the second wave. | | All detected cases will be tested with Ct values. |
| 6 | A typical first wave and a plateaued second wave# | Symptomatic cases have a 25% detection probability during both waves. | | |
| 7 | Two typical waves# | Severe cases have a 37.5% detection probability (1.5 times the baseline), while mild cases have a 20% detection probability (80% of the baseline) during both waves. | Similar detection delays for mild and severe cases. | |

| | | | |
|---|---|---|---|
| 8 | | Symptomatic cases have a 25% detection probability during the first wave;<br>Severe cases have a 37.5% detection probability (1.5 times the baseline), while mild cases have a 20% detection probability (80% of the baseline) during the second wave. | |
| 9 | | Symptomatic cases have a 25% detection probability during the first wave;<br>Severe cases have a 37.5% detection probability (1.5 times the baseline), while mild cases have a 20% detection probability (80% of the baseline) during the second wave, with the detection probability for severe cases further increase during the peak. | |
| 10 | | Symptomatic cases have a 25% detection probability during the first wave;<br>only severe cases have a 25% detection probability during the second wave. | |
| 11 | | Severe cases have a 37.5% detection probability (1.5 times the baseline), while mild cases have a 20% detection probability (80% of the baseline) during both waves. | Longer detection delays for severe cases. |

| | | |
|---|---|---|
| 12 | | Symptomatic cases have a 25% detection probability during the first wave;<br>Severe cases have a 37.5% detection probability (1.5 times the baseline), while mild cases have a 20% detection probability (80% of the baseline) during the second wave. |
| 13 | | Symptomatic cases have a 25% detection probability during the first wave;<br>Severe cases have a 37.5% detection probability (1.5 times the baseline), while mild cases have a 20% detection probability (80% of the baseline) during the second wave, with the detection probability for severe cases further increase during the peak. |
| 14 | | Symptomatic cases have a 25% detection probability during the first wave;<br>only severe cases have a 25% detection probability during the second wave. |

First wave: 0-109 days since the outbreak; second wave: 110-200 days since the outbreak.

### A typical wave is characterized by distinct phases of increase and decrease in case counts, where the $R_t$ rises above 1 during the growth phase and falls below 1 during the decline. In contrast, a plateaued wave due to epidemic characteristics is simulated with $R_t$ fluctuating around 1.

**Table S3. Viral shedding parameters for six real-world pathogens of respiratory viral infections.**

| Pathogen | | Incubation period | | Duration of viral shedding | | Timing of viral peak relative to onset | Quantity of viral peak | |
|---|---|---|---|---|---|---|---|---|
| | | From literature (normal) | After conversion (lognormal) | From literature | From infection (lognormal)* | | From literature | After conversion (normal) |
| SARS-CoV-2 | Alpha | mean=5 and SD=2.3(21) | Mean=4.44 and SD=1.86 | Around 3 days shorter than ancestral strain(6) | Mean=19.56 and SD=1.36 | At onset (similar to non-VOC)(6) | Higher than ancestral ($10^{7.7}$)(6) | Mean=19.95 and SD=4.18 |
| | Delta | mean=4.3 and SD=2.4(21) | Mean=3.75 and SD=2.03 | Around 6 days longer than ancestral strain(6, 9) | Mean=28.84 and SD=1.23 | 1 day before onset(6) | Higher than ancestral ($10^{7.6}$), similar to Alpha(6, 8) | Mean=20.29 and SD=4.18 |
| | Omicron | mean=3.2 and SD=2.2(22) | Mean=2.83 and SD=2.28 | Similar to ancestral strain after onset(5) | Mean=20.45 and SD=1.11 | around 3 days after onset(12-14) | Similar to ancestral(5) | |
| SARS-CoV-1 | | mean=4 and SD=0.2#(23) | Mean=4.00 and SD=1.05 | Around 20 days after onset(10, 24) | Mean=23.98 and SD=1.04 | around 9 days after onset(10, 11) | Trivially higher than ancestral ($1.9\times10^{7}$)(6, 24) | Mean=21.37 and SD=4.18 |
| Influenza A | | mean=1.4 and SD=0.24#(23, 25) | Mean=1.39 and SD=1.16 | Mean=4.8 and SD=0.25#(15) | | 1 day after onset(11) | Trivially higher than ancestral ($1.6\times10^{7}$)(6, 26) | Mean=21.62 and SD=4.18 |

### SD is approximated from CIs (see Supplementary Methods).

* See Supplementary Methods.

#### Supplementary figures

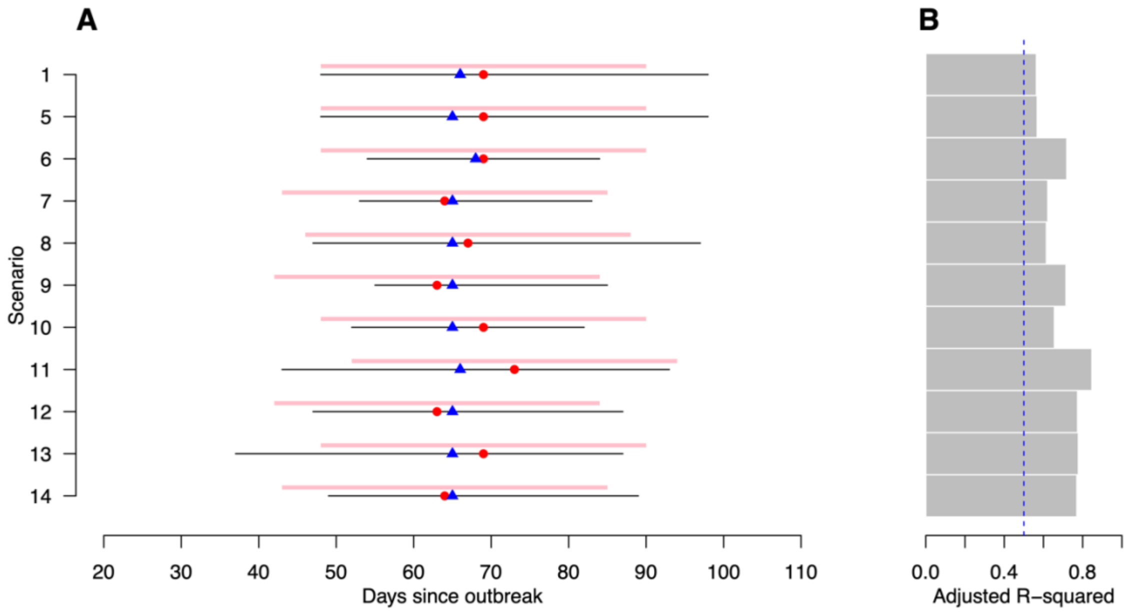

**Fig S1. Summary of best-performing training periods. (A)** time span of best-performing training periods in relation to dates of highest case count and $R_t$ switching points. Each horizontal black line indicates the time period of the best-performing training period (i.e. the time period with the highest adjusted R-squared based on within-sample fit among all alternative training periods in each scenario; see **Supplementary Method**). Red and blue dots indicate the date when highest case count is recorded and the date when $R_t$ changes from above 1 to below 1. Pink bars indicate the time period of 3 weeks before and after the date of highest case count, which is then taken as the generic training period for all scenarios. **(B)** adjusted R-squared of the best-performed training periods. Each horizontal bar indicates the adjusted R-squared for the best-performed training period for each scenario (corresponding to the time period indicated in panel **A**).

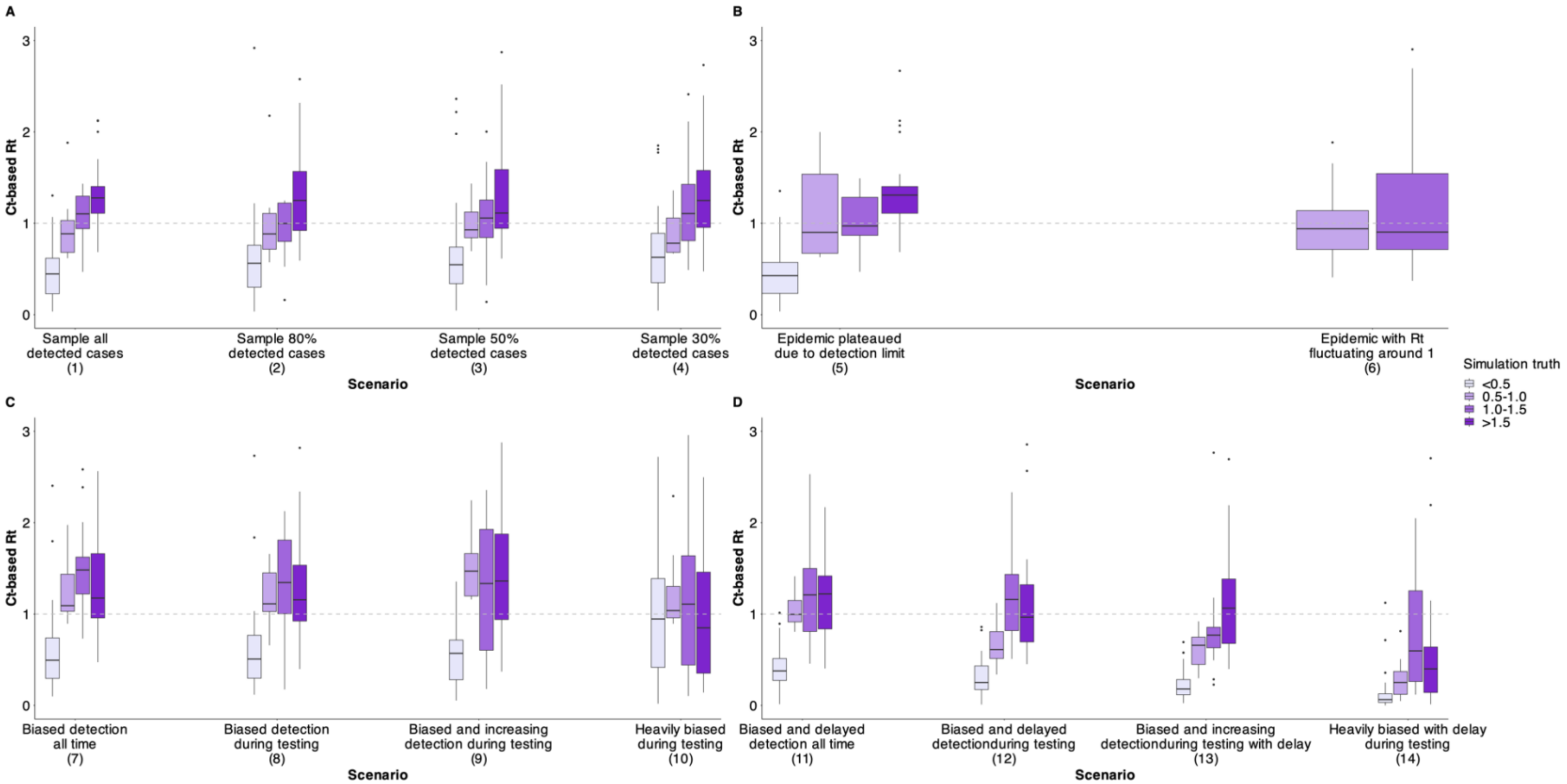

**Fig S2. Correlations between Ct-based $R_t$ and simulation truth during the testing period in scenarios with varying surveillance coverage. (A)** correlations in scenarios with different sampling fractions, **(B)** correlations in scenarios with testing periods plateaued due to different reasons, **(C-D)** correlations in scenarios of biased detections, without **(C)** or with **(D)** further delays for severe cases. Colored boxes indicate the distribution of Ct-based $R_t$ across simulation truth intervals. In scenarios 6, days with $R_t$ between 0.5-1 and 1.0-1.5 are 52 and 39, while days with $R_t$ below 0.5, between 0.5-1, 1-1.5 and over 1.5 in other scenarios are 39, 10, 14 and 28 respectively. Boxes represent the interquartile range (IQR) and median of Ct-based $R_t$, lower whiskers represent the minimum and upper whiskers represent either the maximum or the largest observed values that are within the distance of 1.5 times the IQR of all Ct-based $R_t$ under that simulation truth interval, dots represent values beyond the lower and upper whiskers. Gray dashed line indicates the reference level of Ct-based $R_t$ being 1.

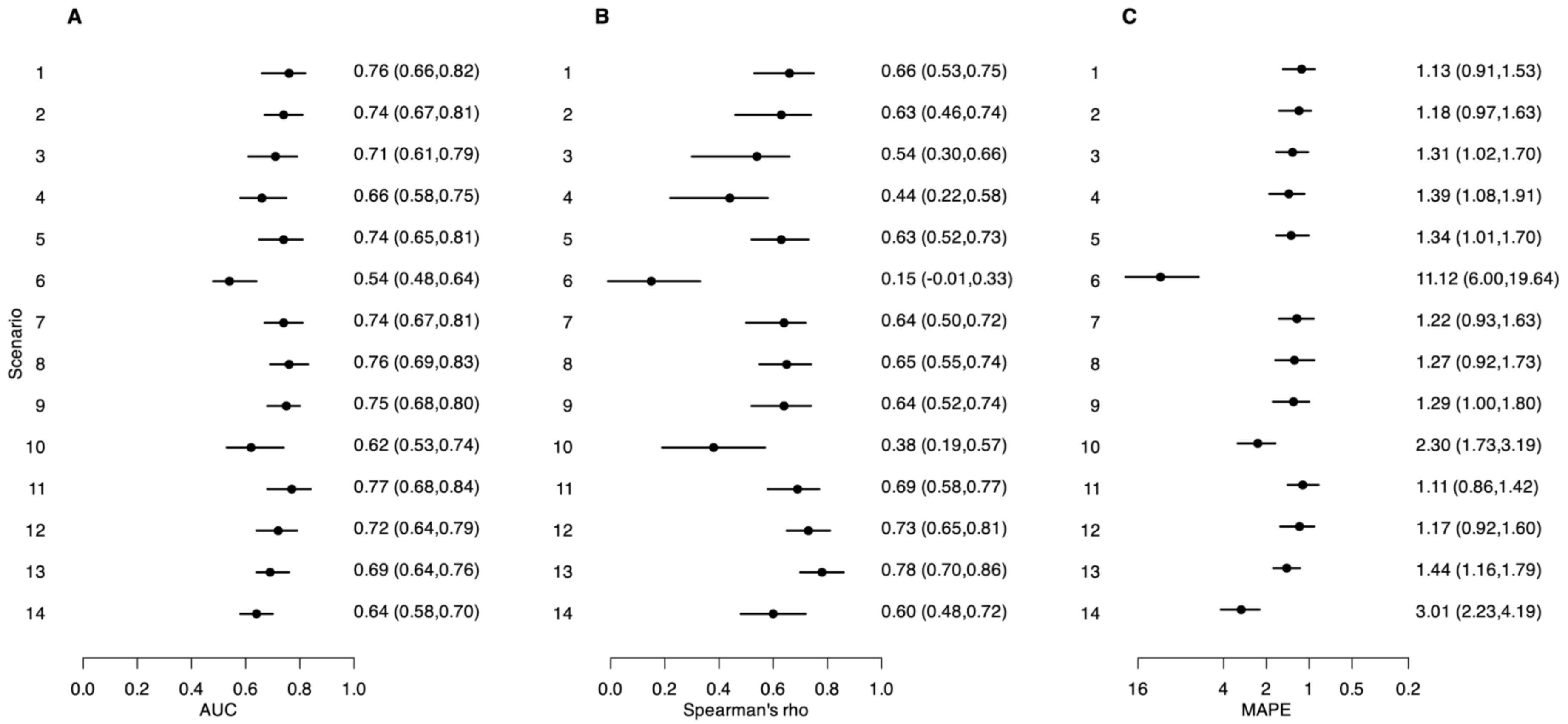

**Fig S3. Evaluation metrics for measuring the estimation accuracy between Ct-based $R_t$ and the simulation truth across simulated surveillance scenarios: (A) AUC, (B) Spearman's ρ, and (C) MAPE.** Points and vertical lines represent median and interval estimates (2.5% and 97.5% percentiles) from 100 bootstrap iterations, with corresponding numeric values shown at right.

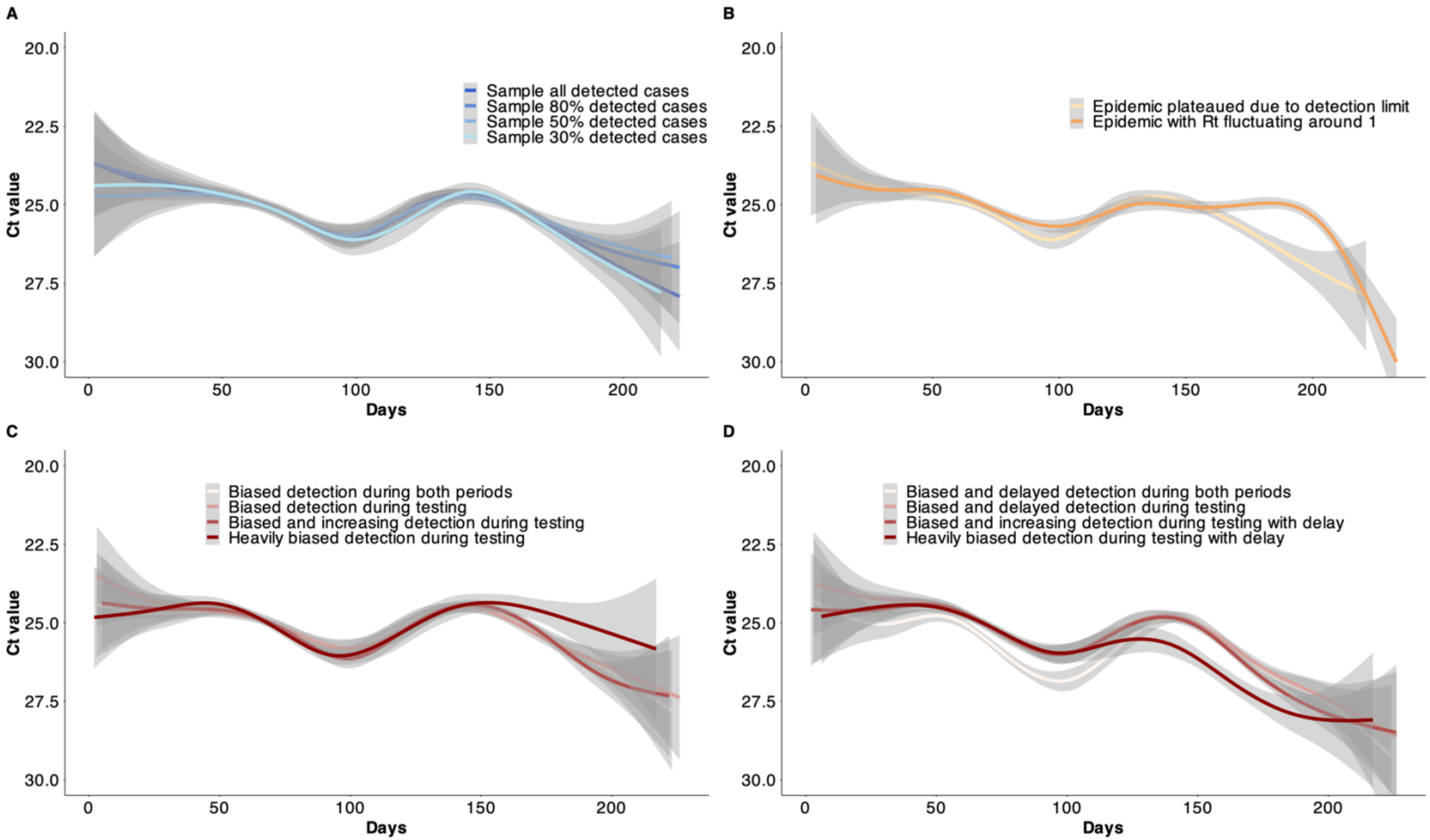

**Fig S4. Temporal variation of population Ct values in scenarios with varying surveillance sensitivity. (A)** temporal variation of population Ct values in scenarios with different sampling fractions, **(B)** temporal variation of population Ct values in scenarios with plateaued testing periods, **(C)** temporal variation of population Ct values in scenarios of biased detections without further delays for severe cases. **(D)** temporal variation of population Ct values in scenarios of biased detections with further delays for severe cases. Lines and shaded areas represent the mean and 95% CIs of smoothing splines for population Ct values over sampling dates in different scenarios, estimated using generalized-additive models.

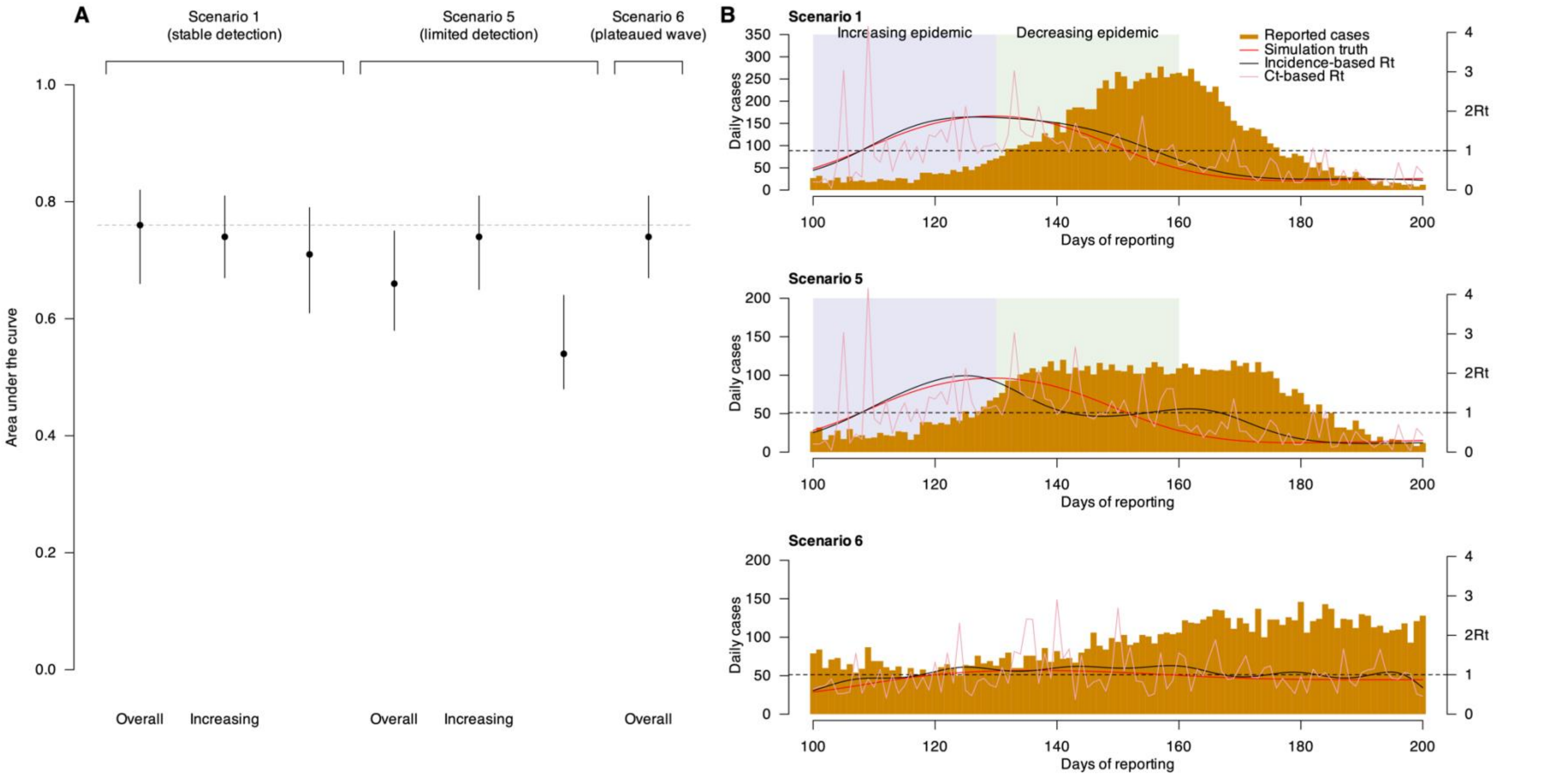

**Fig S5. Estimation accuracy for Ct-based $R_t$ over different epidemic periods in scenarios of stable detection (scenario 1), limited detection (scenario 5) and in a wave with $R_t$ stable around 1 (scenario 6). (A)** estimation accuracy of Ct-based $R_t$ in different time periods, across comparing scenarios. Points and vertical lines show median and interval estimates of the AUC over 100 times of bootstrapping, with the interval estimates taken as the 2.5 and 97.5 percentiles of all 100 estimated values during bootstrapping. **(B)** epidemic curve corresponding to each comparing scenario. Orange bars indicate number of reported cases, and different color lines indicate different estimated $R_t$. Purple and green shaded areas indicate the time period with increasing and decreasing epidemics.

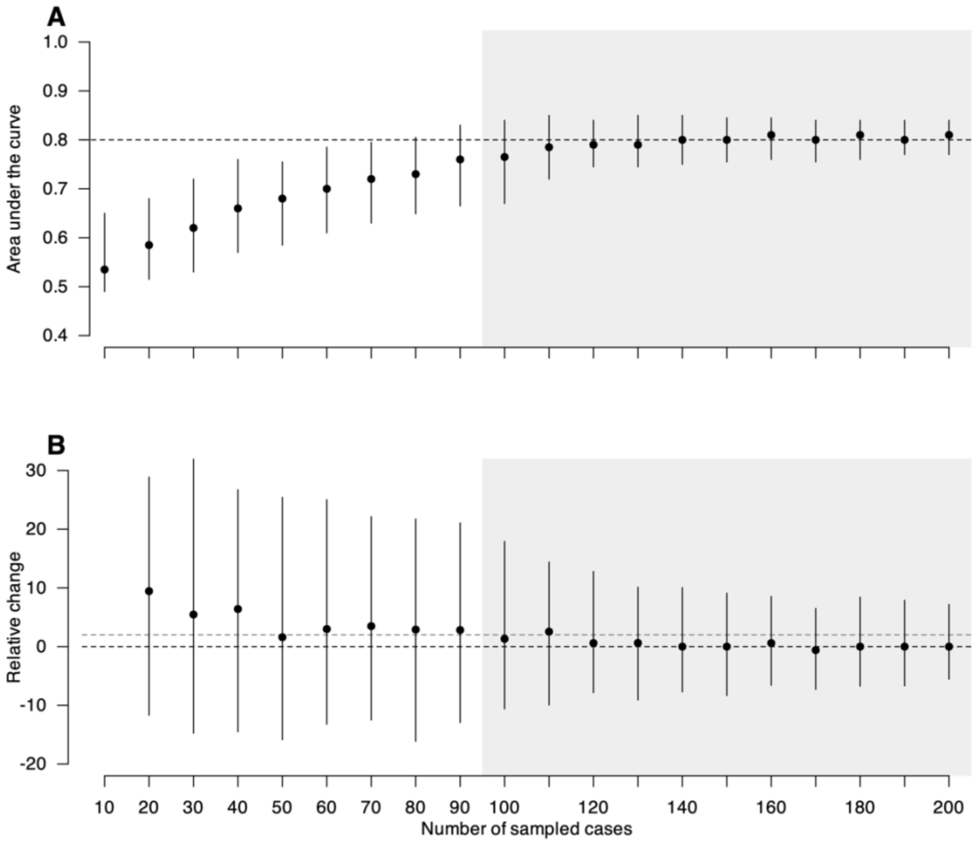

**Fig S6. Absolute values and relative changes in AUC for Ct-based $R_t$ estimated using different available numbers of Ct value samples.** Estimation accuracy in the scenario with stable detection (scenario 1) is used to explore the potential sample count threshold. **(A)** estimation accuracy for Ct-based $R_t$ estimated using different available numbers of Ct value samples. Points and vertical lines show median and interval estimates of the AUC over 100 times of bootstrapping, with the interval estimates taken as the 2.5 and 97.5 percentiles of all 100 estimated values during bootstrapping. Dashed line indicates the AUC when sampling all detected cases as reference. **(B)** relative changes of AUC comparing the AUC estimated using current sample count to the AUC estimated using previous (fewer) sample count (see **Supplementary Methods**). Points and vertical lines show median and the 2.5 and 97.5 percentiles of the relative change of AUC over 100 times of bootstrapping. Black and gray dashed line indicates change of 0% and 2% respectively, as reference. Gray shaded background indicates the situation beyond 100 samples, which we define as marginal improvement.

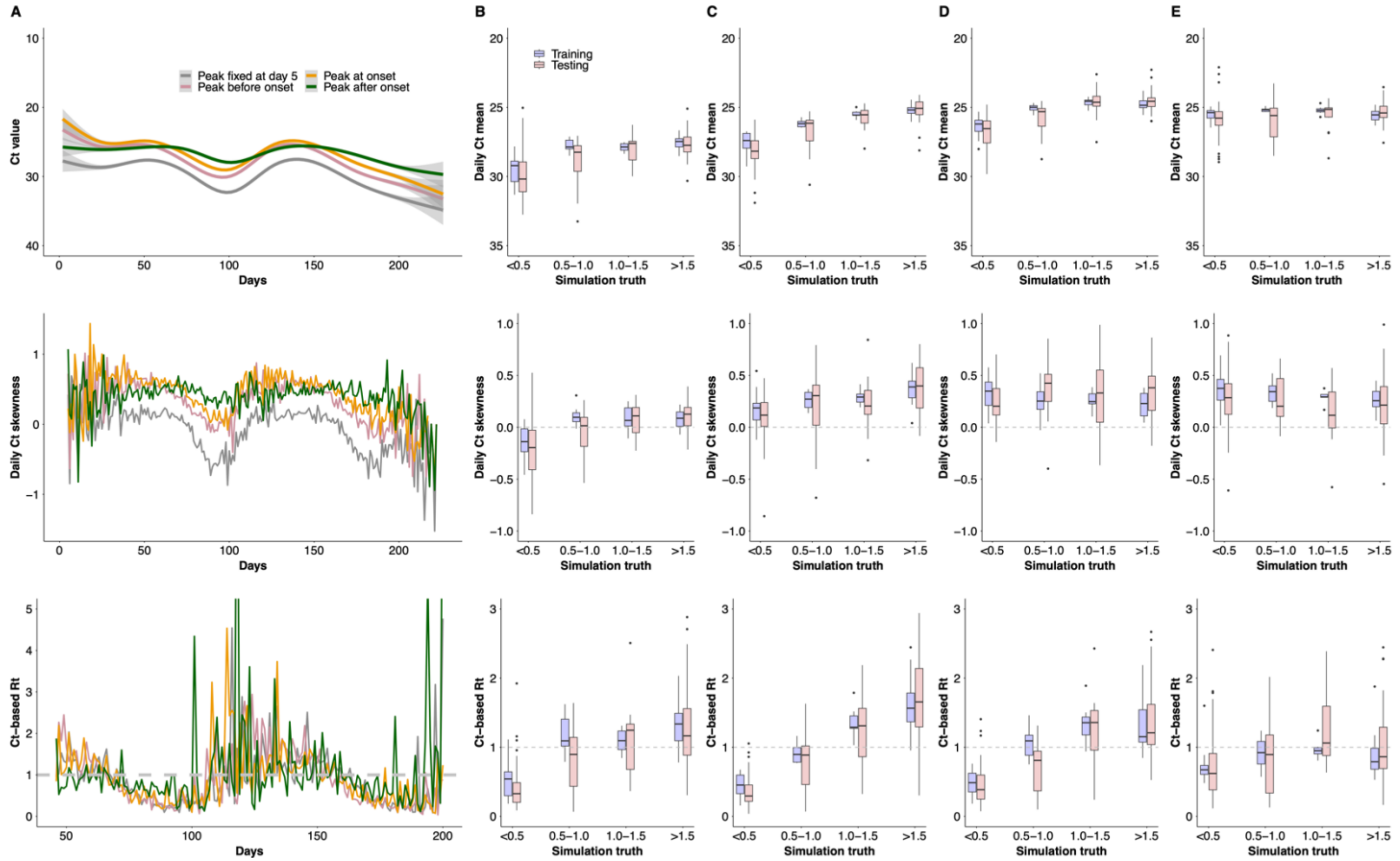

**Fig S7. Temporal variations of population Ct values and estimated Ct-based $R_t$, and their correlation with simulation truth in four example pathogens. (A)** Temporal variations in smoothed Ct values (upper panel), daily Ct skewness (middle panel) and estimated Ct-based $R_t$ (lower panel) for four example pathogens, as denoted in different colors. Smoothed Ct values were estimated from generalized-additive models. **(B-E)** Distributions of daily mean Ct values (upper panels), daily Ct skewness (middle panels) and the estimated Ct-based $R_t$ (lower panels) under simulation truth intervals for four example pathogens respectively. Boxes represent the IQR and median of the distribution under the corresponding simulation truth intervals, lower whiskers represent the minimum and upper whiskers represent either the maximum or the largest observed values that are within the distance of 1.5 times the IQR of all available values under that simulation truth interval, dots represent values beyond the lower and upper whiskers.

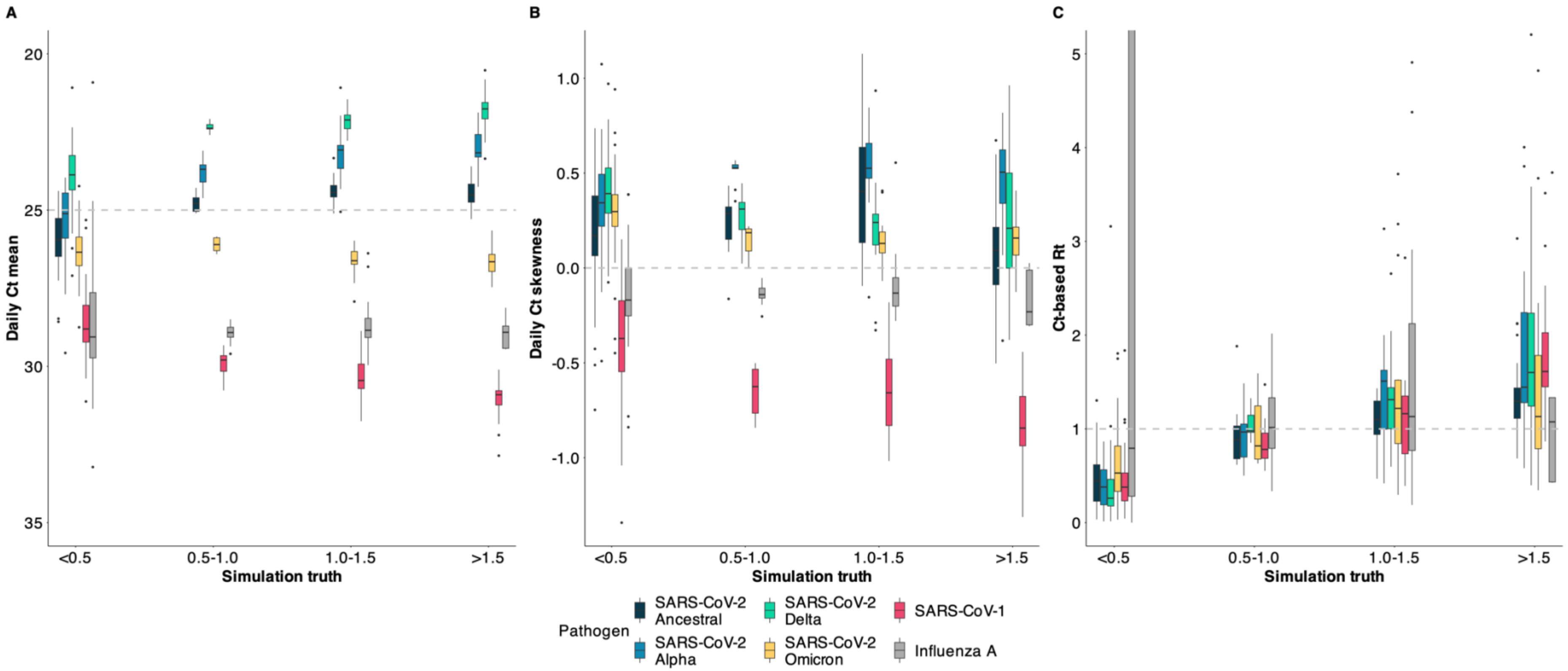

**Fig S8. Correlation between population Ct value distribution or Ct-based $R_t$ in relation to the simulation truth in six real-world pathogens. (A)** Distributions of daily mean Ct values, under simulation truth intervals for six pathogens respectively. **(B)** Distributions of daily Ct skewness under simulation truth intervals for six pathogens respectively. **(C)** Distributions of estimated Ct-based $R_t$ under simulation truth intervals for four example pathogens respectively. Boxes represent the IQR and median of the distribution under the corresponding simulation truth intervals, lower whiskers represent the minimum and upper whiskers represent either the maximum or the largest observed values that are within the distance of 1.5 times the IQR of all available values under that simulation truth interval, dots represent values beyond the lower and upper whiskers.

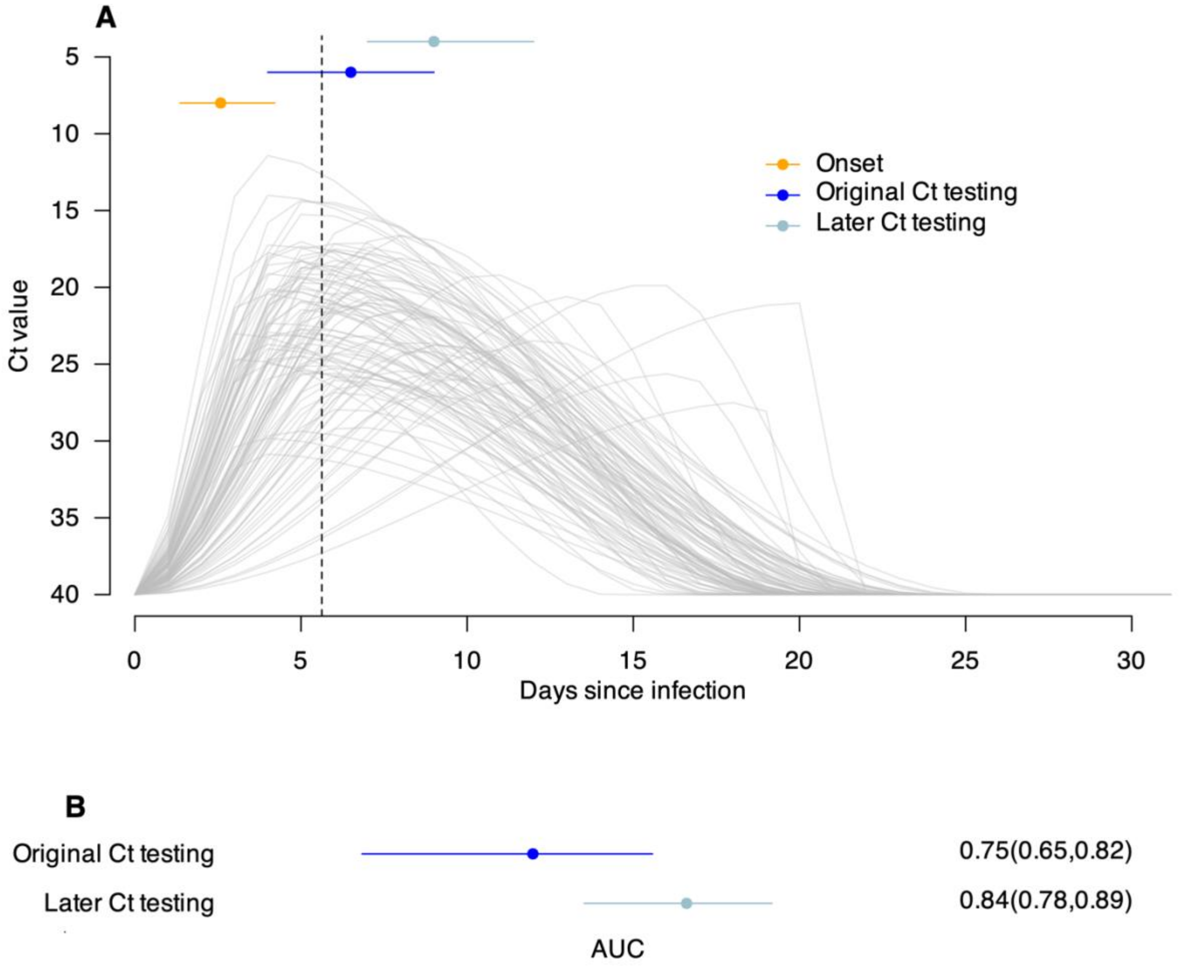

**Fig S9. Impact of Ct testing window on the estimation accuracy of the Omicron scenario. (A)** viral shedding trajectories of 100 randomly selected individuals infected with Omicron. Each gray line indicates the shedding trajectory of an infected individual. The dashed vertical line indicates the median time of viral peak for the 100 individuals. Orange dots and horizontal lines represent the median and IQR of the onset time for these individuals, while dark blue dots and lines represent the median and IQR of their detection time as their original Ct testing time, and light blue dots and lines represent the median and IQR of their Ct testing time if testing is done 2-4 days after case detection. **(B)** estimation accuracies of Ct-based $R_t$ over testing periods when Ct testing was done at detection (original Ct testing) or 2-4 days after detection (later Ct testing) in the Omicron scenario. Dots and horizontal lines represent median and interval estimates of the AUC over 100 times of bootstrapping, with the interval estimates taken as the 2.5 and 97.5 percentiles of all 100 estimated AUC values during bootstrapping.
